# E-norms at Ten: What a Decade Taught the Method About Deriving Reference Values from Patient Data

**DOI:** 10.64898/2026.08.30.26361424

**Authors:** Joe F Jabre

## Abstract

**Introduction/Aims:** The e-norms method derives reference values from a laboratory’s own mixed patient data, without recruiting healthy volunteers. A decade of use exposed three things the original description did not address: a dependence on how finely measurements are recorded, a minimum sample size below which derived limits are systematically too permissive, and an extension of the method to engineered composite features. This work characterizes the first two and describes a corrected plateau rule.

**Methods:** Twenty-six sensory amplitude datasets (18 to 3,301 measurements), three sensory conduction velocity datasets, nine median-nerve datasets and a 2,000-measurement peroneal file were analyzed. Amplitudes were progressively rounded; velocities were made finer by adding noise of 0.05 m/s. Subsamples of 50 to 750 were drawn from thirteen large datasets on raw, logarithmic and square-root scales. The corrected rule was validated against expert visual plateau selection and against the plateau proportions reported in 2015, and limit stability was estimated by resampling.

**Results:** At n = 50, amplitudes as recorded returned 0.28 of the full-sample limit with 73.1% failing; rounded to whole numbers they returned 1.03 with none failing. Integer velocities returned 1.03 with no failures, but failed in all 300 draws once noise was added. Derived limits were systematically too low below 250 to 300 measurements. The corrected rule returned 1.00 with 0.1% failures and left integer velocities essentially unchanged.

**Discussion:** The dependence is intrinsic to locating the plateau from gaps between sorted values. Obtaining 250 normal studies means accumulating 530 to 1,250, depending on a laboratory’s normal rate.

## 1. Introduction

The e-norms method was developed and was being used in our laboratory since the late nineties as a way to develop our EMG laboratory’s own normal values from diagnostic studies we performed on patients referred to us for electrodiagnostic studies. At first, we used it on EMG and Nerve Conductions studies, then in jitter analysis and blood serum electrolytes, the method being data agnostic. Submitting our work describing the method for publication at the time was very challenging given that few healthcare providers were willing to embrace the concept of deriving normal values from patient data. That changed after a serendipitous encounter with a pediatric clinical neurophysiologist who had developed a new single fiber EMG technique to study young children referred to rule out neuromuscular transmission disorders called

SPACE [1], and needed to develop normal values for these studies where none were available in the literature. This chance encounter led to the method’s first publication in 2015 in the Journal of Clinical Neurophysiology [2], [3]. Use of the method spread steadily thereafter, driven by a simple fact: the interpretation of any diagnostic study depends on the reference values applied to it, and for most laboratories, the collection of their own requires the recruitment of a healthy cohort to derive those from, obtaining ethical approval for the study, the performance of these studies, and their analysis — an undertaking of months or years that few laboratories can fund or staff with. The practical result being that most laboratories routinely use values collected elsewhere, from a different cohort, on different equipment, by differently trained examiners, and sometimes obtained decades earlier.

This is the assumption that the formal theory of reference values starts from as well [4] — that a person recruited as normal is, and the results collected from them are, considered “a-priori” normal. The alternative we developed with the e-norms method was to look at how measurements behave *after* a laboratory’s routine clinical studies have been done, and to identify which of them cluster in the way normal values do. We refer to this as an “a-posteriori” approach to developing normal values, since they are derived after a study is completed, and it is what the e-norms method implements. Another downside of the a-priori approach is that it is not merely difficult but in certain instances unavailable, and they are worth stating concretely because they are the situations that motivated this work.

As an example, some tests have no volunteers. A relatively infrequent case in point is worth mentioning here, needle examination of the anal sphincter (sphincter ani) is used in the investigation of fecal incontinence, and needle examination of the tongue (genioglossus) in the investigation of amyotrophic lateral sclerosis. Very few volunteers without symptoms would consent to either, and consequently no healthy-cohort reference values exist for them. Yet among patients referred for those studies, 58 of 103 sphincter ani examinations and 60 of 90 genioglossus examinations were normal (unpublished data). The needle exam of these muscles is performed for a clinical reason; the normal values were already there among patients who were examined for these symptoms, some of whom turned out to have a negative study.

But more importantly, some cohorts such as pediatrics or the elderly cannot be recruited at the time they are needed. In pediatrics, reference values for nerve conduction in infants in the first weeks of life are very infrequently available in the literature, and the reason is not that such infants are never studied — it is that a mother a few weeks after delivery is very unlikely to bring her newborn in to volunteer.

And some tests are too new for any cohort to exist. Procedures and measurements now being developed with the help of artificial intelligence have no normative literature to borrow from and no healthy population that has yet been measured, because there has been no time to measure one. In that situation the a-posteriori approach is not the convenient option; it is the only one.

The method itself is simple. E-norms sorts a laboratory’s measurements in an ascending order, identifies the flat middle segment of the resulting curve — the plateau, where consecutive values differ least — and derives reference limits from the values lying within it.

In the decade since its first publication, the e-norms method has been applied to pediatric and geriatric nerve conduction and electromyography [5], [6], [7], [8], evoked potentials, neuromuscular ultrasound [9], ophthalmic biometry [10], orthopedic measurement [11], inflammatory markers [12] and body mass index [13] among many others too numerous to cite.

Since the e-norms method first appeared in 2015, three methods using its same underpinnings for deriving reference values from mixed patient datasets have appeared [14], [15], [16], resting on the same underlying observation, an idea first proposed by Hoffmann in 1963 [17]. These methods have since been compared with one another, and against reference limits derived in the traditional way [18], [19], [20]. Acceptance of the concept has grown alongside them, and its use has spread well beyond clinical neurophysiology, triggered by our development of an e-norms web app in 2017 (https://enorms.com) where users can upload their data on the web and calculate their own normal values in it. To date, more than 1.5 million measurements from over 20 countries have been analyzed with the e-norms method web implementation [21].

A decade of the method’s use has of course exposed things the original description did not address. In late 2025 we therefore undertook a systematic review of our own work with the method, and this paper reports the three findings that came out of it.

The first is that the plateau step depends on how finely the measurements were recorded — a dependence that had gone unnoticed; it is invisible in data recorded as whole numbers and destructive in data recorded to decimals.

The second is that sample size acts on the method through the number of repeated values it accumulates, which sets a minimum below which the derived limit is not merely unstable but systematically too permissive.

The third is that the method extends beyond directly measured variables to engineered composite features, such as in its recent application by this author to EEG, an application that opens it to signals that cannot be sorted in their native form. We refer to this recently published e-norms application as e-norms+ [22]. That work is reported separately and is discussed rather than presented here.

The aim of this work is to characterize these dependencies, to describe a corrected plateau rule that removes the first of them, and to set out what a laboratory needs in order to use the method reliably.

To do this, the method itself was treated as the object of study rather than as a tool. The material analyzed was reference-value data already held from clinical neurophysiology laboratories: sensory nerve action potential amplitudes (SNAP), sensory conduction velocities, and a motor conduction velocity dataset.

During this work, it became clear to us that the dependence of the method on recording precision was not anticipated; it emerged from the behavior of these datasets during analysis and was then tested deliberately, and that is the sequence the Methods section below will follow.

## 2. Methods

Two properties of the data were implicated by the initial analysis: how finely the measurements had been recorded, and how many of them there were. The experiments described below alter each of these deliberately, and observe what happens to the plateau as a result. Amplitudes were made progressively coarser. Conduction velocities were made finer by the addition of noise far below the resolution of any instrument in clinical use. Subsamples of increasing size were drawn from the largest files, and each subsample was analyzed three times over: once on the measurements as recorded, once after taking their logarithms, and once after taking their square roots. The corrected plateau rule, its two independent validations, an estimate of how stable a derived limit is, and a calculation of how much data a laboratory must accumulate all follow from what those experiments showed.

### 2.1 Data

All data were nerve conduction measurements collected in the course of routine clinical work and provided by the contributing laboratories for methodological review and analysis. No healthy volunteers were recruited, and no measurement was made for the purpose of this study. The measurements were anonymized before they were transferred and contain no patient identifiers, contributing laboratory or provider information or any other personal health record information; each contributing laboratory had obtained institutional review board approval for the collection of the data at source.

These datasets were not assembled to test any particular property of the method. They were the material available, and the questions this paper addresses arose from their behavior during analysis rather than preceding it. What made them suitable, once those questions had been raised, is that they span two recording conventions and a wide range of sizes: amplitudes are recorded to one or two decimal places while conduction velocities are recorded as whole numbers, and the largest amplitude files hold enough measurements to be subsampled repeatedly. Four groups of data were used.

They were:

Sensory amplitudes: Twenty-six independent datasets of sensory nerve action potential amplitude, from the median nerve (digits II and III) and the sural nerve, recorded in children and grouped into age bands, containing between 18 and 3,301 measurements each, most between 300 and 1,400.

Sensory conduction velocities: Three datasets: median (n = 388), ulnar (n = 387) and radial (n = 84). The median nerve set: Nine datasets, orthodromic and antidromic, digits II and III, amplitude and conduction velocity, of 49 to 193 measurements each. These served as small-sample examples throughout.

A motor reference file: Two thousand peroneal motor conduction velocity measurements, used throughout as the large, stable comparator.

Thirty sensory amplitude files were supplied and twenty-six were analyzed; the remainder were duplicate submissions of the same data or empty files. No dataset was excluded on the basis of its results, and no value was removed from any dataset apart from a single physiologically impossible conduction velocity, removed as a transcription error before any analysis.

Not every analysis used every dataset. The number of datasets entering each analysis is stated with the result.

### 2.2 The plateau rule as published in 2015

The e-norms method sorts the variable measurements of a mixed laboratory population into ascending order and plots them [2]. The resulting curve has three parts: a steep lower segment, a flat middle segment, and a steep upper segment. The flat middle segment — the plateau — is taken to hold the values that are normal for that laboratory (Figure 1), and descriptive statistics computed on it give the reference range: the plateau mean plus and minus two standard deviations, with the values at the ends of the plateau reported separately as the observed minimum and maximum.

**Figure 1:**
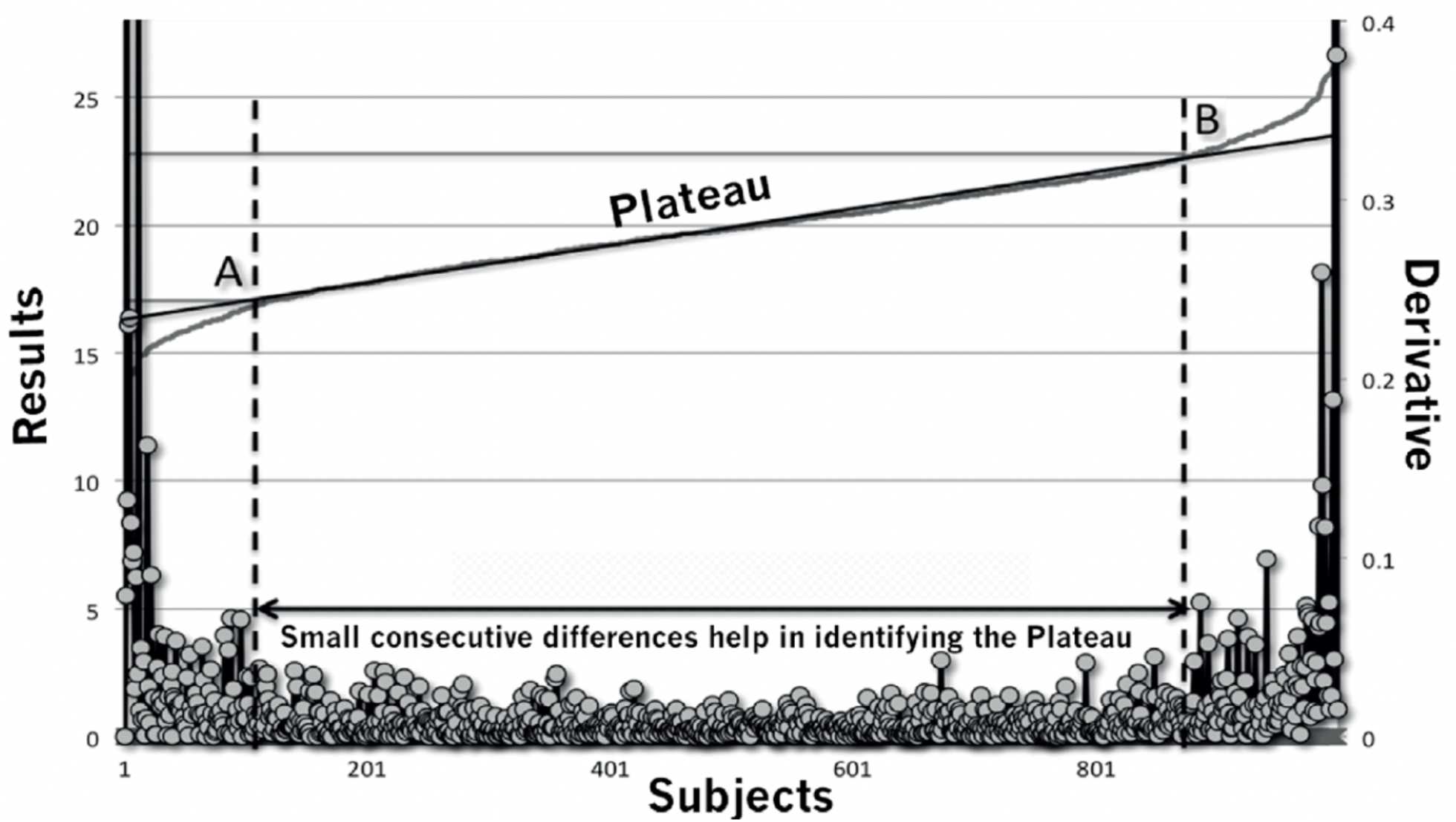
The e-norms principle, illustrated with single fiber EMG jitter data. Measurements are sorted in ascending order and plotted against rank (left axis, Results). The curve has a steep lower segment, a flat middle segment and a steep upper segment; the flat middle segment — the plateau — is taken to hold the values that are normal for that laboratory. The scatter along the bottom shows the first-order differences between consecutive sorted values (right axis, Derivative). The plateau corresponds to the region where those differences are smallest. Its ends, marked A and B, were located visually by the user in the procedure as originally published. (Adapted from Jabre JF, Pitt MC, Deeb J, Chui KKH. E-norms: a method to extrapolate reference values from a laboratory population. J Clin Neurophysiol 2015;32(3):265–270).

The plateau is located from the first-order differences between consecutive sorted values. The published rule identifies the region where those differences are smallest and change most slowly. In the implementation that has been used since, a group is formed of the values lying at the smallest consecutive difference. The mean and standard deviation of that group define a window extending one standard deviation either side of its mean. Every value in the dataset falling inside that window constitutes the plateau, and the reference limits are the plateau mean plus and minus two standard deviations. Plateau identification was originally performed visually and by hand; it was automated in 2024 with a Python program developed by the author [23].

### 2.3 Experiment 1: recording precision

The dependence of the plateau step on how finely the data are recorded was tested in both directions on the same datasets.

Amplitudes were made coarser under three conditions: as recorded, rounded to one decimal place, and rounded to whole numbers. Conduction velocities, which were recorded as whole numbers, were made finer by adding Gaussian noise with a standard deviation of 0.05 m/s — an amount well below the accuracy of any instrument in clinical use and clinically invisible.

Every condition was run at a sample size of 50, with 300 random draws, and each draw’s lower reference limit was expressed as a ratio to the lower limit obtained from that dataset’s own full sample. A draw was recorded as a failure when the procedure returned no usable limit. The proportion of duplicate values and the number of distinct values were measured for each condition.

### 2.4 Experiment 2: sample size, and the scale on which the data are analyzed

Thirteen amplitude datasets with at least 800 measurements were used as parent populations. Three hundred subsamples were drawn at each of seven sizes — 50, 100, 150, 200, 300, 500 and 750 — and each subsample’s lower limit was again expressed as a ratio to the parent dataset’s own full-sample limit. Because the subsamples are drawn from the parent, each preserves the parent’s own proportion of abnormal studies; the effect of sample size is therefore isolated from the effect of contamination.

The same procedure was repeated with each subsample analyzed three ways — on the measurements as recorded, on their logarithms, and on their square roots — to establish whether transforming the data helps, and under what conditions.

One limitation of this design should be stated plainly here. The reference in every case is the dataset’s own full-sample answer, not an externally established truth. These experiments therefore measure convergence and stability, not accuracy.

### 2.5 The corrected plateau rule

The behavior described in Section 2.3 was traced back to the implementation, where it proved to arise from two defects that are in fact one mechanism.

First, the consecutive differences were computed in a way that placed an artificial value of zero at the first row. Zero is smaller than any real difference, so the smallest difference was always this artificial one, and the lowest value in the dataset was always drawn into the group.

Second, where no two measurements in a dataset were equal, that artificial first entry was the only member of the group. The standard deviation of a single value is undefined, so every statistic downstream became blank — without an error, and without any indication that the result was missing rather than computed.

The correction keeps the published rule exactly as it stands: collect the values lying at the smallest consecutive difference. It adds one condition. If fewer than ten values lie at that smallest difference, the group is widened to take in the values at the next-smallest difference as well, and widening continues until at least ten values are in the group. Ten is the smallest number from which a mean and standard deviation can be computed with any stability, and Section 3.4 shows the result is not sensitive to that choice. The artificial leading difference was removed. The second stage is unchanged: the group’s mean and standard deviation define the one-standard-deviation window from which the plateau, and hence the reference limits, are computed.

An alternative correction was tried first and rejected. Taking a fixed percentage of the smallest differences fails where repeated values are abundant: thousands of differences are then exactly zero, the choice among them is settled by sort order, and selection is biased toward the lowest values. On the peroneal file this produced a lower limit of 34.35 against 39.75 from the published rule.

The corrected rule was applied to the experiments of Sections 2.3 and 2.4, and to 39 datasets in a direct comparison of the limits obtained before and after.

### 2.6 Validation of the corrected rule

Two validations were performed, independent of each other and of the experiments above.

The first compares the automatic result against expert visual selection. The dataset on which the two rules disagree most was plotted in the e-norms web implementation, the plateau was selected visually on the sorted curve as it would be in ordinary use, a judgment shown to be reproducible across observers [24], and the reference limits were then recomputed from the rank range selected.

The second compares the proportion of each dataset falling inside the plateau against the proportions reported in the original 2015 paper, which were 45% in a pediatric cohort and 62% in an adult cohort. This proportion was computed for 28 datasets under both the original and the corrected rule.

### 2.7 Stability of the derived limit

The lower limit of each of 38 datasets was recomputed on 400 resampled versions of that dataset, and the width of the resulting 95% interval was taken as the measure of how much the limit depends on which particular patients happened to be included.

Resampling with replacement is not straightforwardly valid for a method built on repeated values, and the reason is the subject of this paper. Drawing with replacement manufactures repeated values that were never in the data: the duplicate rate rises from 46.7% to about 61% in the peroneal file, and from 29.1% to about 49.5% in the largest amplitude dataset. The resampled datasets are therefore systematically more tied than their parent, and the limits computed from them are displaced. On the peroneal file, under the corrected rule, the naive interval of 37.85 to 38.97 does not contain the limit computed from the real data, 39.98.

The width of the resampled distribution is nonetheless informative even where its center is not. Spread was therefore measured by resampling and then reported centered on the limit computed from the real data. Both the corrected and the uncorrected intervals were retained.

### 2.8 How much data a laboratory must collect

Because e-norms extracts normal values from studies that have already been performed, the practical question facing a laboratory is not how many healthy volunteers to recruit but how many routine studies to accumulate. This reverses the question the literature usually asks — the level of contamination at which a method breaks — into one a laboratory can act on.

The number of studies to collect is the target number of normal studies divided by the laboratory’s normal rate, the proportion of its studies reported as normal. A 2014 survey of 200 electromyographers worldwide, with 47 respondents (Jabre, unpublished), gives a mean normal rate of 47%, with a range from 20% to 80%.

### 2.9 Software and verification

Analyses were performed in Python 3 with NumPy, pandas and SciPy. Before use, the corrected rule was reimplemented independently from its written description, without reference to the working code, and the two agreed to within 1 × 10⁻⁹ on every dataset tested.

### 2.10 AI-assisted analysis

Computational analyses were carried out through human–AI collaboration using Claude (Anthropic). The investigator specified the questions, the methodological framework and the interpretation of results; the AI implemented the computations, performed the statistical calculations and drafted code that was reviewed and accepted by the investigator before it was applied to data. All methodological decisions, interpretations and conclusions remained the responsibility of the investigator.

## 3. Results

### 3.1 The plateau step depends on how finely the data are recorded

The plateau is located from the smallest differences between sorted values, so repeated values are what drive it. Conduction velocities, recorded as whole numbers, had duplicate rates of 84.5% to 94.3% and held only 13 to 34 distinct values, so a sample of 50 still contained 31 to 38 repeats. SNAP amplitudes, recorded to decimals, had duplicate rates of 1.2% to 29.1%, so a sample of 50 contained very few repeats, and on the least granular files almost none. Where repeats run out, the group defining the plateau collapses to a single value and the result is blank.

Coarsening the amplitudes repaired them completely, and adding clinically invisible noise to the velocities destroyed the method outright (Table 1).

**Table 1.**
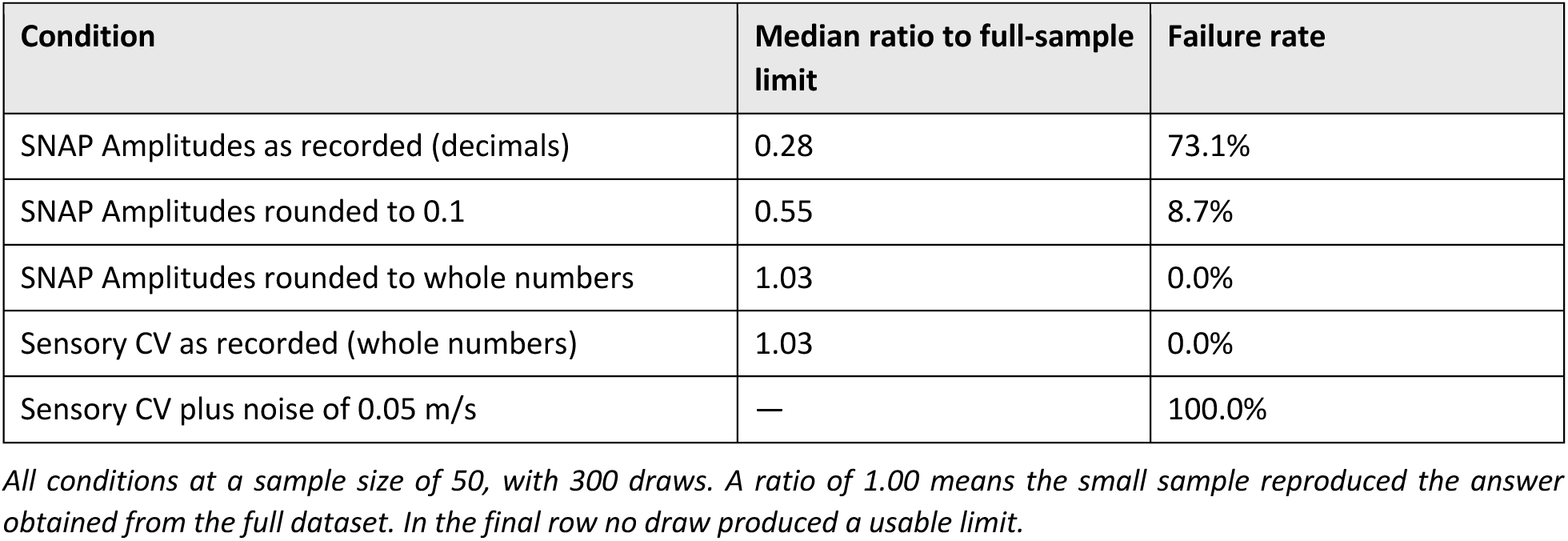
Effect of recording precision on the plateau step.

| Condition | Median ratio to full-sample limit | Failure rate |
| --- | --- | --- |
| SNAP Amplitudes as recorded (decimals) | 0.28 | 73.1% |
| SNAP Amplitudes rounded to 0.1 | 0.55 | 8.7% |
| SNAP Amplitudes rounded to whole numbers | 1.03 | 0.0% |
| Sensory CV as recorded (whole numbers) | 1.03 | 0.0% |
| Sensory CV plus noise of 0.05 m/s | — | 100.0% |

*All conditions at a sample size of 50, with 300 draws. A ratio of 1.00 means the small sample reproduced the answer obtained from the full dataset. In the final row no draw produced a usable limit.*

Sample size, in this method, has only ever mattered as a proxy for the number of repeated values accumulated.

### 3.2 Sample size acts through the number of repeated values

Failure and bias are two separate phenomena, and the second is the more important (Table 2). Outright failure clears by 300 measurements. But below that size the limit is not merely unstable — it is systematically far too low, which is the permissive direction: an abnormal study is called normal.

**Table 2.** Lower limit obtained from subsamples, as a ratio to the full-sample limit.

| Sample size | Raw: median ratio | Raw: failure rate | Log scale: median ratio |
| --- | --- | --- | --- |
| 50 | 0.31 | 72.7% | 0.44 |
| 100 | 0.33 | 27.1% | 0.47 |
| 150 | 0.48 | 6.1% | 0.54 |
| 200 | 0.65 | 0.9% | 0.62 |
| 300 | 0.83 | 0.0% | 0.75 |
| 500 | 0.95 | 0.0% | 0.89 |
| 750 | 0.99 | 0.0% | 0.97 |

Convergence to within 5% of the full-sample answer requires about 500 measurements, and to within 1% about 750.

### 3.3 The scale of analysis: the recommendation is conditional on sample size

Below about 200 measurements the logarithmic scale is markedly less biased and tighter than the raw scale. At 200 the two cross. Above it the raw scale converges faster (Table 2). The square-root scale sits between the two at every size.

The recommendation is therefore conditional on sample size and never universal. This resolves an apparent disagreement in the literature: a logarithmic transformation was chosen on very large datasets and a square-root transformation on a dataset of a few hundred, and both choices were reasonable under the condition each faced. The condition itself has not previously been stated.

Shape alone is the wrong trigger for transforming. Across the 26 amplitude datasets every skew was positive, from +0.16 to +1.67, yet not one produced a negative lower limit on the raw scale. A rule that transformed on the basis of skew would have transformed all 26 unnecessarily. On large datasets the three scales agree closely — 13.81, 14.06 and 14.20 on the largest, of 3,301 measurements — and they diverge only on small, highly skewed ones, where the sample size is itself the problem.

### 3.4 The corrected rule

The corrected rule repairs the amplitude failure, leaves data rich in repeated values untouched, and makes the method insensitive to recording precision (Table 3). The last row is the important one: it is the property the precision experiment showed to be missing.

**Table 3.** Behavior of the original and corrected rules.

| Case | Original rule | Corrected rule |
| --- | --- | --- |
| Peroneal motor CV file, full sample | 39.75 – 49.50 | 39.98 – 49.29 |
| SNAP Amplitudes at n = 50 | 0.31 of full-sample limit; 72.7% failed | 1.00 of full-sample limit; 0.1% failed |
| Sensory CV at n = 50 | 1.03; 0.0% failed | 1.04; 0.0% failed |
| Sensory CV plus 0.05 m/s noise | 100% failed | 1.06 – 1.12; 0.0% failed |

On the peroneal file the group defining the plateau already held 933 values, so the widening condition never engaged; the small movement in its limits comes entirely from removing the artificial leading difference, which had been dragging the dataset’s lowest value into the group. The result is not finely tuned to the minimum chosen: minima of 5, 10 and 20 gave median ratios of 1.04, 1.00 and 0.98 on the amplitude data, with failure rates of 0.4%, 0.1% and 0.0%.

Across 39 datasets compared directly, 18 were essentially unchanged, moving by one unit or less, and 21 changed materially. The largest upward movement was +9.89 µV, on a sural amplitude dataset of 164 measurements whose lower limit rose from 4.09 to 13.98; the largest downward movement was −2.15 m/s, on a small median antidromic velocity dataset. Every dataset of 700 measurements or more was stable, and two of the three sensory velocity datasets recorded as whole numbers were unchanged, the third moving by 2.05 m/s.

One consequence should be stated rather than left for a reader to find. On the smallest datasets, those poorest in repeated values, the corrected band can be narrow: one median sensory velocity dataset of 67 measurements moved from 51.78–67.74 to 59.43–64.32, a range under 5 m/s. The correction stops silent failure and removes the downward bias, but it does not make a small dataset trustworthy. Datasets below roughly 250 measurements remain unreliable whatever rule is applied to them.

### 3.5 First validation: the corrected rule agrees with visual selection

The sural amplitude dataset of 164 measurements, on which the two rules disagree most, was examined visually. Judged from the sorted curve, the plateau should begin at about rank 35 to 40; the automatic selection under the original rule had begun it at rank 10. Recomputing from rank 35 gives a lower limit of 14.95 µV, and from rank 40, 16.18 µV. The corrected rule returns 13.98 µV automatically. The original rule returned 4.09 µV.

The corrected rule therefore lands within about two microvolts of the expert judgment the method was built to reproduce [24], and the original was wrong by a factor of three on the same data.

At the upper end the agreement is less close: the corrected rule gives 34.07 µV against about 38 µV from the visual selections. The corrected plateau window on this dataset holds 81 of 164 measurements, or 49%, against 70% for the plateau as drawn by hand. The automatic plateau is narrower at both ends, and the upper limit follows from that rather than from a separate defect — a point Section 3.6 bears on directly.

### 3.6 Second validation: the corrected rule reproduces the plateau proportions of 2015

The 2015 paper reported that the plateau held 45% of a pediatric cohort and 62% of an adult cohort [2]. Across 28 amplitude and velocity datasets the corrected rule gives a median plateau proportion of 54.3%, ranging from 42.0% to 73.6%. The original rule gave a median of 55.5% over a wider range, from 32.9% to 73.6% — and the 32.9% floor was the sural dataset that failed.

The corrected rule therefore sits inside the experience reported in the original paper and its range is tighter. It also puts the hand-drawn plateau of Section 3.5, at 70%, above the original paper’s own reported experience, which suggests that the visual selection there was over-inclusive at the lower end rather than that the automatic result was too narrow.

### 3.7 Stability of the derived limit

Across 38 datasets the average width of the 95% interval on the lower limit was 3.54 units where the dataset held 250 measurements or more, and 12.20 units where it held fewer — a 3.4-fold difference (Table 4). This reproduces the 250 to 300 threshold by a technique entirely different from the subsampling of Section 3.2, and independently of the published recommendations that also converge on that range.

**Table 4.** Width of the 95% interval on the lower reference limit.

| Dataset size | Average width of the 95% interval |
| --- | --- |
| 250 measurements or more | 3.54 units |
| Fewer than 250 measurements | 12.20 units |

*Four hundred resampled replicates per dataset, 38 datasets. Width is expressed in the units of the measurement. The interval is centered on the limit computed from the real data, for the reason given in Section 2.7.*

The extremes make the point concretely. The tightest intervals came from the largest datasets: 1.32 units on an amplitude dataset of 3,301 measurements and 1.12 m/s on the peroneal file of 2,000. The widest was 27.42 µV on an amplitude dataset of 60 measurements — an interval wider than the reference range it was meant to qualify. Six datasets were flagged as unstable on that criterion, the interval covering more than half the reference range, and all six held between 49 and 87 measurements.

### 3.8 How many studies a laboratory must collect

Combining the normal rates reported by 47 responding electromyographers with the sample sizes established above gives a table a laboratory can use directly (Table 5). At the survey mean of 47%, obtaining 250 normal studies means accumulating 532; at a normal rate of 20% it means accumulating 1,250.

**Table 5.** Studies a laboratory must accumulate to obtain a given number of normal studies.

| Target normal studies | 20% | 30% | 40% | 47% | 60% |
| --- | --- | --- | --- | --- | --- |
| 100 | 500 | 334 | 250 | 213 | 167 |
| 250 | 1,250 | 834 | 625 | 532 | 417 |
| 300 | 1,500 | 1,000 | 750 | 639 | 500 |
| 500 | 2,500 | 1,667 | 1,250 | 1,064 | 834 |

*Column headings are the laboratory's normal rate, the proportion of its studies reported as normal. The 2014 survey of 200 electromyographers worldwide, with 47 respondents, gives a mean of 47%, ranging from 20% to 80%. A target of 100 normal studies is included for completeness only; the results above indicate that fewer than 250 does not give a dependable limit.*

## 4. Discussion

### 4.1 The plateau, and its dependence on recording precision

In this section, we begin with the plateau, the mainstay of the e-norms method. Everything e-norms produces comes from the values lying within it, and every question about the method’s reliability is ultimately a question about whether the plateau was located and identified correctly. What a decade of use of this method has revealed is that the step which locates it is sensitive to something we had not thought to examine: not the physiology, not the size of the dataset, but how finely the measurements happened to be written down.

The plateau is found by looking for the region where consecutive sorted values differ least. That construction has a consequence which becomes obvious once stated: it depends entirely on values being repeated, or nearly repeated, within the data. Conduction velocities recorded as whole numbers repeat constantly. Amplitudes recorded to two decimal places almost never do. The same procedure applied to the two behaves quite differently, and the difference is a property of the recording convention rather than of the measurement.

The direction of the effect is counterintuitive. Recording more precisely makes the method work less well, and adding noise far below the resolution of any instrument in clinical use was enough to stop it entirely. This is not a defect peculiar to one implementation. Any procedure that locates the plateau from the gaps between sorted values inherits the same dependence.

There is convergent evidence for this in the literature on related methods. The extrapolated reference values (E-Ref) procedure originally located its reference point from the angle of the sorted curve — also a construction built on the gaps between neighboring values. Tied values arising from integer recording were reported to make that curve stepwise and its angle plot uneven, producing spurious peaks [25], and successive versions of the procedure moved away from angle measurement, first to smoothing and curve fitting, then to slope, and finally to a construction based on tangents drawn from the mean and standard deviation of the whole dataset [26]. Ties have therefore been described as a pitfall for one family of plateau locators and are, in the method described here, the very thing the plateau step relies upon. Both observations follow from the same mechanism.

The corrected rule removes the dependence without changing what the method is. It keeps the published definition — collect the values lying at the smallest consecutive difference — and adds a single condition: if that group is too small to compute statistics from, widen it to the next-smallest difference until it is large enough. Data rich in repeated values are unaffected, because the condition never engages. Data poor in repeated values are repaired.

It follows from the mechanism, rather than from any result reported here, that a procedure estimating the density of the measurements directly would not have this dependence at all — not because it performs better, but because it does not read gaps. A preliminary test was not sufficient to establish it, which makes the approach insufficiently tested rather than untested. No claim is made for it here.

### 4.2 How much data is needed, and on what scale it should be analyzed

We turn next from the procedure to the data it is given. Two questions have been asked separately in the literature and are treated separately in practice: how many measurements a laboratory needs before the derived limit can be trusted, and whether those measurements should be analyzed as recorded or transformed first. Where datasets are small, or distributions markedly skewed, different authors have recommended different transformations — logarithmic in some work, square root in others — without stating the conditions under which each applies. The results reported here indicate that the two questions are really one, because which scale performs better depends on how much data there is.

Taking the transformation question first: below roughly 200 measurements the logarithmic scale is less biased and tighter than the raw scale; above it the raw scale converges faster. A recommendation made on a dataset of several hundred and a recommendation made on datasets of several thousand will therefore disagree, and neither is wrong. What has been missing is the statement that the recommendation is conditional on sample size at all.

A second point follows. Skew alone is the wrong trigger for transforming. Every one of the 26 amplitude datasets examined here was positively skewed, and not one produced a negative lower reference limit on the raw scale. A rule that transformed on the basis of distributional shape would have transformed all 26 without need. The practical trigger is not the shape of the data but the size of the dataset.

Turning to size itself, three independent lines of evidence converge on the same figure. Subsampling from large parent datasets shows the derived limit approaching the full-sample answer at around 250 to 300 measurements, with convergence to within 5% at about 500. Resampling shows the 95% interval on the lower limit widening more than threefold below 250. And published recommendations for related methods arrive in the same region from different directions [14], [15], [16], [19], [26].

Below that size the problem is not instability but direction. Small samples do not scatter around the right answer; they return limits that are systematically too low, which is the permissive direction — an abnormal study called normal. A laboratory deriving a reference limit from 100 measurements is not merely working with an imprecise number. It is working with a number that is probably too generous, and nothing in the output says so.

This is where the two questions join. The scale of analysis matters most in precisely the range where the sample is too small to give a dependable limit in the first place. Above 200 to 300 measurements the choice of scale becomes secondary, and on the largest datasets the three scales agree closely enough that it hardly matters. A laboratory with enough data does not have a transformation problem; a laboratory that has a transformation problem is being told something about its sample size.

The practical question this raises is not how many normal studies are needed but how many studies must be accumulated to obtain them, and that depends on the proportion of a laboratory’s work reported as normal. Among 47 responding electromyographers to an unpublished survey, that proportion ranged from 20% to 80% with a mean of 47%. At the mean, 250 normal studies means accumulating about 530; at 20%, about 1,250. For most laboratories this is a matter of months of ordinary work rather than a research project, which is the point of deriving reference values from work already done.

### 4.3 E-norms+: extension to engineered composite features

The principle of the e-norms method is that a laboratory should more appropriately compare a patient’s measurements against normal values derived from its own patient cohorts — recorded on its own equipment, with its own electrodes, filter settings and technique, and in the population it actually serves. That principle began in electromyography and nerve conduction studies and has since been carried into ophthalmic biometry, neuromuscular ultrasound, orthopedic measurement, inflammatory markers and body mass index among many others. A recent extension of the e-norms method we now refer to as e-norms+ [22], has now carried it into the field of electroencephalography through the development of a composite stability metric constructed for data that are continuous and multidimensional rather than the single values to which every earlier application of the method was confined. This section sets out what that extension requires, because it also bears directly on the correction described in Section 4.1.

Every application listed above operates on a directly measured quantity — an amplitude, a velocity, a latency, an axial length, a body mass index. Each is a single number per subject, and each can be sorted. Signals that are not single numbers have therefore been outside the method’s reach. A multichannel electroencephalogram is a set of continuous waveforms in which the same voltage may be normal or abnormal depending on its frequency content and its context, and which cannot meaningfully be sorted in its native form.

The extension is to interpose a constructed variable. Rather than sorting the signal, one computes from it a composite feature that aggregates several of its characteristics into a single dimensionless number per unit of time, and then applies e-norms to that. The requirement is not that the feature be a physiological quantity, only that it be one number per unit, that it vary in the direction of interest, and that its normal values cluster more tightly than its abnormal ones — the clustering behavior the method has always depended on. In work reported separately [22], a composite of first-derivative dynamics, spectral entropy, variance and line length, computed per two-second epoch across 23 channels, satisfied these conditions well enough that e-norms applied to it recovered patient-specific baselines from seizure-free recordings and identified seizures without any labeled training data.

This extension, which may be termed e-norms+ [22], changes what kind of data the method can address. It also carries the dependency described in Section 4.1 with it. A constructed feature is a continuous quantity with no recording convention to coarsen it, so it is exactly the case in which repeated values are scarce and the corrected plateau rule matters most. The two findings of this paper are therefore linked: the reach into composite features is what makes precision-independence necessary rather than merely tidy.

What the extension does not do is guarantee that any composite feature will work. The construction of the feature is a modeling decision made before e-norms is applied, and a poorly chosen feature will produce a clean plateau around a quantity of no clinical meaning. The method identifies the population that clusters; it does not verify that the thing being clustered is worth measuring.

### 4.4 What deriving reference values from patient data with mixed datasets is actually for

Having described what the method depends on and how far it now reaches, it is worth returning to the question that motivated its development in the first place: why derive reference values from patients at all, when the conventional alternative exists. Reference values are ordinarily obtained by deciding in advance that a particular group of people is healthy, measuring them, and treating the result as normal. E-norms works the other way around. It takes measurements already made, without any prior judgement about who is or was healthy, and identifies which of them behave as normal values are deemed to behave. The first is an a-priori approach; the second is an a-posteriori one.

The recurring objection to the a-posteriori approach is that patients are not normal subjects. That objection assumes normal subjects are available and can be identified in advance. Three experiences over the past decade suggest the assumption is weaker than it appears.

The first is a matter of timing. In 1996, before the e-norms method had been published, it was applied to 200 anonymized samples from a hospital chemistry laboratory. Chloride, Potassium, Sodium and Creatinine came back close to the values the hospital was using derived from normal values available at the time in the literature. Total cholesterol and fasting glucose did not: the e-norms method put the upper limit of cholesterol below 183 mg/dL where the laboratory used 225, and glucose at 101 mg/dL where the laboratory used 120. At the time this looked like a failure of the method, and nearly ended the work. In the two decades since, accepted values have moved toward the lower figures [27], [28]. The explanation is not that the method was clever but that the a-priori cohort was defined by the absence of current symptoms, and a 25-year-old carrying a cholesterol of 225 mg/dL has no symptoms, or a glucose of 120 mg/dL — until the age of 50 or 60. A healthy cohort assembled today is blind to disease that has not yet declared itself.

The second is published and can be checked. Applying the method to 34,384 body mass index measurements gave a healthy range of 22.7 to 30.6 [13]. The upper figure sits close to the accepted threshold for obesity. The lower figure is far above the WHO and CDC threshold of 18.5 — and it corresponds closely to the point, in a cohort study of 3.6 million adults, at which all-cause mortality begins to rise [29]. Once again, the e-norms method’s number, rather than the official one, matched where the harm actually starts.

The third is a limitation rather than a vindication, and it belongs here for that reason. In the same work, healthy BMI in the 80-to-98 age group came out lower than actuarial studies suggested. One explanation is that chronic pathology in the very old biases conventional estimates upward. Another is survivor bias: those who reached that age were probably healthier, and likely carried a lower BMI earlier in life. On that reading, the method was not describing a hypothetical healthy population aged over 80 but a healthy sub-population of survivors.

These three sit together. The a-priori approach requires knowing in advance who is healthy, and that knowledge can be unavailable because the disease has not declared itself, because no cohort can exist yet, or because the cohort that does exist has been shaped by who survived to be in it. The a-posteriori approach describes the population that is actually present. That is its strength in the first two cases and its boundary in the third, and both should be stated.

### 4.5 Limitations

A method’s limits are part of its specification. Knowing where the e-norms method stops being dependable is what allows it to be used with confidence everywhere else, and each limit below marks work that would extend the method rather than a reason to distrust it.

The first limit is one of size, and it qualifies the correction itself. The corrected rule stops silent failure and removes a downward bias, but it does not make a small dataset trustworthy. On the smallest, tie-poor files the resulting band can be implausibly narrow, and datasets below roughly 250 measurements remain unreliable whatever rule is applied to them. The correction makes a bad answer visible; it does not make it good.

The second limit concerns what these experiments can establish. The subsampling and resampling analyses use each dataset’s own full-sample answer as their reference, so they measure convergence and stability rather than accuracy against an external standard. They cannot establish that the full-sample answer is itself correct — that would require a matched healthy cohort, which is the thing the method exists to avoid needing. Comparison against mortality data, as in the body mass index work, is one way around this and is worth pursuing for other measurements.

The third limit concerns the data analyzed here. They come from a small number of contributing laboratories and are weighted toward sensory studies in pediatric age bands. Whether the precision threshold and the sample-size minimum hold in the same form for motor studies, for other measurement types, and across a wider range of laboratories is not established by these data, and a multi-laboratory replication would settle it.

The fourth is a question this work raises without answering. The reference limits are reported as the plateau mean plus and minus two standard deviations — a symmetric interval applied to distributions that are frequently skewed. Whether a symmetric band is the right summary of a plateau is separable from everything addressed here, and is left open.

The last concerns the alternative discussed in Section 4.1. The density-based approach has had only preliminary testing. Establishing whether it is a practical replacement would require bandwidth sensitivity analysis, a sweep across measurement types, and a sample-size comparison against the rule described here. Until that is done it remains a direction indicated by the mechanism rather than a result.

## 5. Conclusion

The e-norms method was proposed as a way for a laboratory to recover reference values from work it had already done. A decade of use across neurophysiology, ophthalmology, orthopedics, chemistry and anthropometry among others has confirmed that it does so, and has also shown what it depends on.

The quick answer is, it depends on repeated values. That dependence was invisible for as long as the method was applied to quantities recorded as whole numbers, and it becomes destructive as measurements are recorded more finely — the direction in which instrumentation is moving. The corrected plateau rule removes the dependence while leaving the published definition intact, and agrees both with expert visual selection and with the plateau proportions reported in the original description.

It also depends on sample size, and does so through the number of repeated values a dataset accumulates rather than through sample size as such. Below roughly 250 to 300 measurements the derived limit is systematically too permissive, and the output gives no indication of it. Expressed in terms a laboratory can act on, obtaining 250 normal studies means accumulating between about 530 and 1,250, depending on how much of that laboratory’s work is reported as normal.

And e-norms reaches further than single measurements. Applied to engineered composite features, the e-norms method extends to signals that cannot be sorted in their native form — an extension that also makes the precision correction necessary rather than optional, since constructed features have no recording convention to coarsen them.

But what has not changed is the premise: normal values cluster tightly, while abnormal ones scatter. Tolstoy put the same asymmetry more memorably in Anna Karenina when he wrote that all happy families are alike, each unhappy family is unhappy in its own way [30]. Reference values are present in work that has already been done, on the equipment that will be used to interpret them, in the population that will be tested. The three cases that opened this paper — the tests nobody volunteers for, the cohorts that cannot be recruited in time, and the measurements too new for any healthy population to have been assembled — are not edge cases in which an a-posteriori approach is a reasonable compromise. They are cases in which it is the only approach available.

## Ethics and patient consent statement

This study involved secondary analysis of anonymized nerve conduction data provided by contributing laboratories for the purpose of methodological review and analysis. The data contained no patient identifiers and no laboratory or institutional information, and the contributing laboratories obtained institutional review board approval for the collection of the data at source. As this work involved only retrospective analysis of previously collected, anonymized data, no additional ethical approval or patient consent was required.

## Data availability statement

The data analyzed in this study were provided by contributing laboratories for the purpose of methodological review and are not publicly available. The plateau rule as originally published (Section 2.2), the corrected rule (Section 2.5), and the experiments performed on the data (Sections 2.3, 2.4, 2.6 and 2.7) are specified in sufficient detail to enable reimplementation and independent testing on other datasets.

## Contributions

This work was not performed as part of the author’s academic affiliation with Tufts University School of Medicine. Joe F Jabre is the sole author and was responsible for all aspects of this work including conceptualization, e-norms methodology development, data analysis, validation, interpretation of results, and manuscript preparation. AI collaboration is detailed in the Declaration of generative AI below.

## Declaration of generative AI and AI-assisted technologies in the manuscript preparation process

During the preparation of this work the author used Claude (Anthropic) for computational analysis, statistical calculation, data processing, code development, and for drafting and editing sections of the manuscript. All methodological decisions, interpretations of results, and conclusions were the sole responsibility of the author, who developed the e-norms method, specified all methodological requirements, reviewed and validated every analysis, provided the clinical interpretation, and made all evidence-based decisions. AI served as an analytical tool and collaborator, not as an autonomous decision-maker. After using this tool, the author reviewed and edited the content as needed and takes full responsibility for the content of the published article.

